# A precise score for the regular monitoring of COVID-19 patients condition validated within the first two waves of the pandemic

**DOI:** 10.1101/2021.02.09.21249859

**Authors:** Evgeny A. Bakin, Oksana V. Stanevich, Vassily A. Belash, Anastasia A. Belash, Galina A. Savateeveva, Veronika A. Bokinova, Natalia A. Arsentieva, Ludmila F. Sayenko, Evgeny A. Korobenkov, Dmitry A. Lioznov, Areg A. Totolian, Yury S. Polushin, Alexander N. Kulikov

## Abstract

**Purpose:** The sudden outbreak of COVID-19 pandemic has shown that the medical community needs an accurate and interpretable aggregated score not only for an outcome prediction but also for a daily patient’s condition assessment. Due to a continuously changing pandemic landscape, robustness becomes a crucial additional requirement for the score.

**Patients and methods:** In this research, real-world data collected within the first two waves of the COVID-19 pandemic was used. The first wave data (1349 cases collected from 27.04.2020 to 03.08.2020) was used as a training set for the score development, while the second wave data (1453 cases collected from 01.11.2020 to 19.01.2021) was used as a validating set. For all the available patients’ features we tested their association with an outcome using robust linear regression. Statistically significant features were taken to the further analysis for each of which their partial sensitivity, specificity and promptness were estimated. The sensitivity and the specificity were further combined into a feature informativeness index.

**Results:** The developed score was derived as a weighted sum of the following 9 features showed the best trade-off between informativeness and promptness: APTT (> 42 sec, 4 points), CRP (> 146 mg/L, 3 points), D-dimer (> 2149 mkg/L, 4 points), Glucose (> 9 mmol/L, 4 points), Hemoglobin (< 115 g/L, 3 points), Lymphocytes (< 0,7*10^9/L, 3 points), Total protein (< 61 g/L, 6 points), Urea (> 11 mmol/L, 5 points) and WBC (> 13,5*10^9/L, 4 points). Thus, the proposed score ranges between 0 and 36 points. Internal and temporal validation showed that sensitivity and specificity over 90% may be achieved with an expected prediction range >7 days. Moreover, we demonstrated a high robustness of the score to the varying peculiarities of the pandemic. For the additional simplicity of application we split the full range of the score into five grades delimited with 9, 14, 19 and 24 points which determine expected death:discharge odds 1:100, 1:25, 1:5 and 1:1 correspondingly.

**Conclusions:** An extensive application of the score within the second wave of the COVID-19 pandemic showed its potential for the optimization of patients management as well as improvement of medical staff attentiveness during high workload stress. The transparent structure of the score, as well as tractable cut-off bounds, simplified its implementation into clinical practice.

## Introduction

As of February 2021, there are over 100 million confirmed cases of SARS-CoV-2 infection worldwide [1]. In addition to the avalanche-like increase in infections, we see a significant number of deaths – more than 2 million, which is about 2% of the number of cases. In some regions one can see even higher mortality rates: for example, in Italy the mortality rate is currently 3.4%, in the UK – 2.8%.

Due to a frequent uncontrollable course of the infection, many medical workers are wondering about the possibility of creating an effective and flexible system for assessing the severity of patients’ conditions. Such assessment systems are necessary both for an accurate prediction of a treatment results, and for determination of indications for the use of certain therapy regimens that may have multidirectional effects. Summarizing the experience of managing patients with COVID-19, the researchers came to the conclusion about the different power of various existing scales for adverse outcomes prediction (SOFA, qSOFA, SAPS III, APACHE II) [2]–[4]. Recently, special attention has been paid to the tools for assessing the risk of a lethal outcome based on machine learning algorithms [5], [6].

According to the literature, several indicators are the most interesting for predicting a lethal outcome: aspartate aminotransferase [7], conjugated bilirubin [8], creatinine, urea [9], procalcitonin [10], CRP [11] and lymphocytes [12]. These indicators are useful for death prediction both separately and in combination as was shown with use of automatic feature selection tools based on such machine learning algorithms as logistic regression, SVM, random forest, etc. [13]

However, in spite of all the advantages, the application of these tools in a broad clinical practice remains very limited. We consider the following main reasons for it.

1. An evaluation of some components of the scores may be complicated due to a high cost or the complexity of a measurement procedure. For example, the respiratory index included in SOFA score can be calculated only by means of artery blood taking for an acid-base balance assessment.
2. Usually, in the published papers devoted to prognostic scores development, the evaluation of a patient state is performed only once (e.g. at the moment of admission into a hospital or ICU). However, the application of a score for a patient regular monitoring is another important use case (e.g. for a treatment strategy optimization) which requires an accurate examination of its temporal characteristics such as prediction range.
3. Since 2019, the pandemic landscape was continuously evolving: the new paradigms of treatment changed the old ones, the more dangerous viral strains appeared and widely spread. This makes it necessary to perform a regular validation of predicting approaches and their adjustment if required.
4. Despite the approaches based on state-of-art machine learning algorithms demonstrate a high accuracy (up to 90%), they frequently have a structure too complex for a direct interpretation by a doctor. This complicates their acceptance in the medical community.

### Objective

In this research we aim to develop and validate a novel prognostic score, having a transparent structure and suitable for an everyday evaluation of a patient condition. As an additional requirement we stated the score robustness, cheapness and the wide availability of laboratory tests included in the score.

## Patients and methods

### Study design and participants

In this research we used real-world data collected within the first two waves of COVID-19 pandemic at St. Petersburg State Pavlov Medical University. The first wave data (collected 27.04.2020 - 03.08.2020) was used as a training set for the score development, while the second wave data (collected 01.11.2020 - 19.01.2021) was used as a validating set. For both cohorts, a SARS-CoV-2 infection was proved using a PCR method. Before analysis we excluded patients with a short follow-up period who died or were successfully discharged within the first 2 days of hospitalization. Thus, mostly patients with a moderate or severe initial illness status were accepted to this research, while patients with a mild or critical status were exclude [14]. The patients with a loss of follow-up were excluded from this study as well. The overall study profile is given in Figure 1.

**Figure 1.**
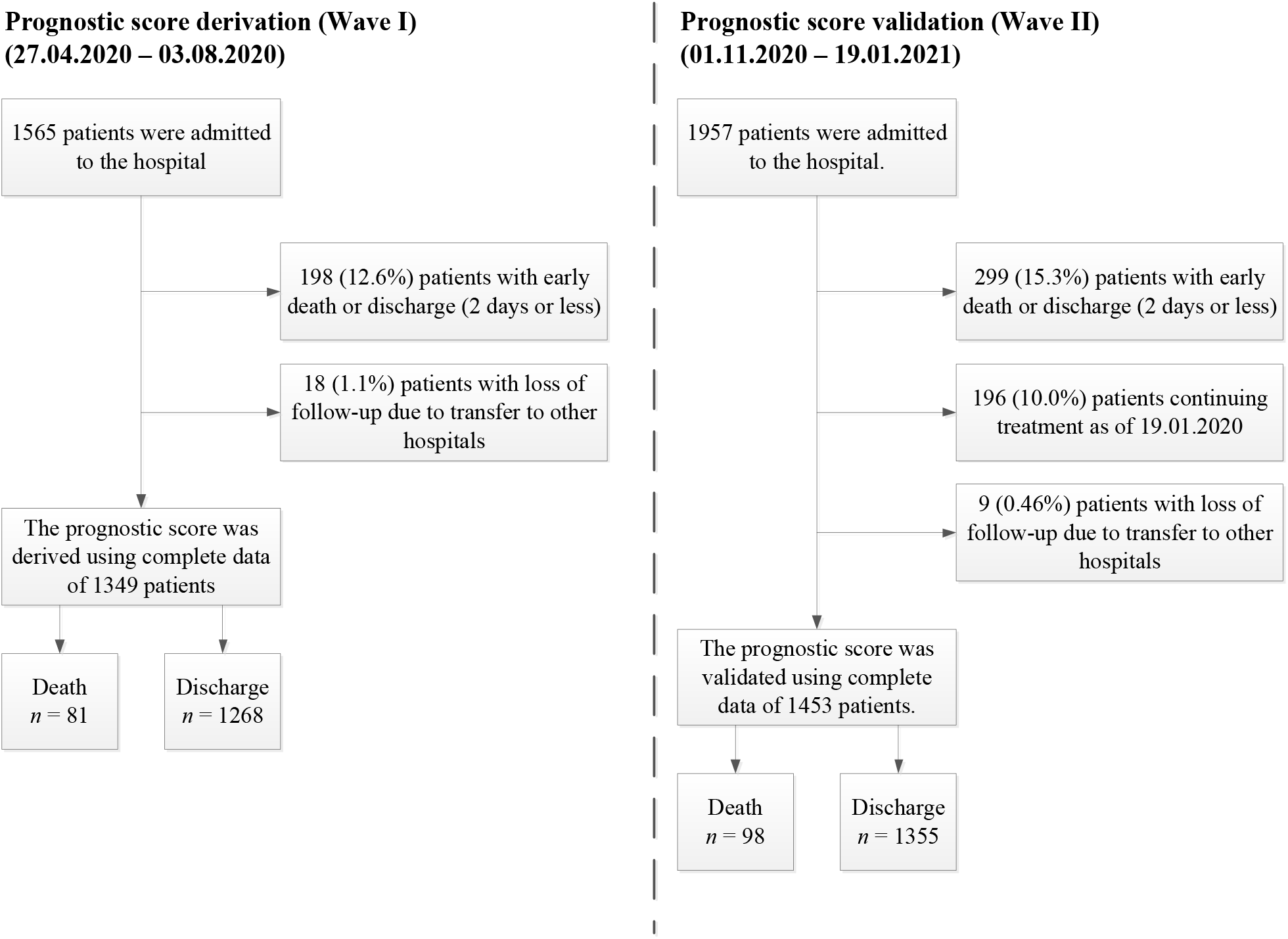
Study profile

**Figure 1.**
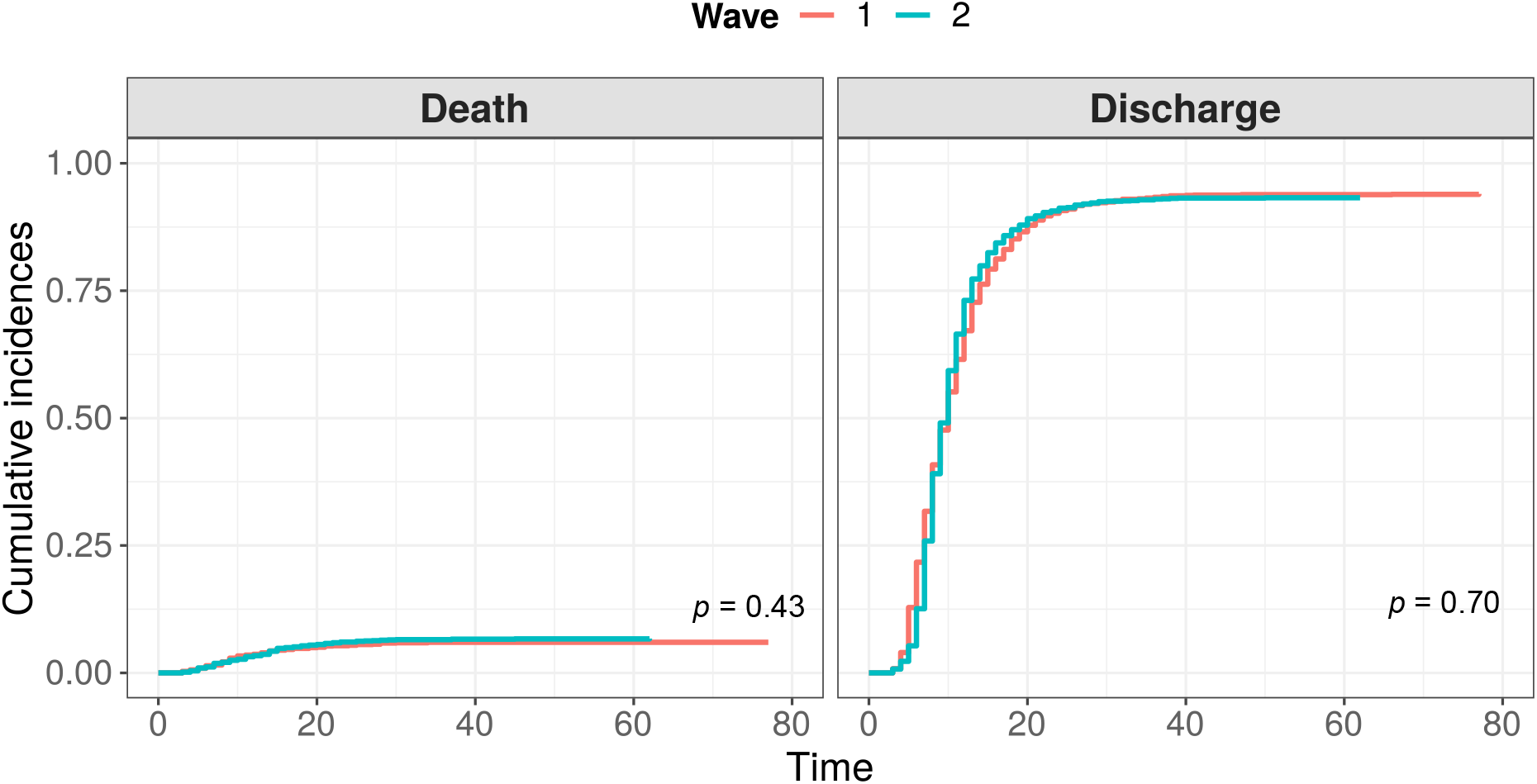
Comparison of cumulative distribution functions for events “Death” and “Discharge” between two waves of COVID-19

### Initial features set

In this research we used the following initial features set routinely gathered from hospital patients:

– *antropometry*: age, sex, BMI;
– *blood differential test*: total white blood cells (WBC), lymphocytes, neutrophils, monocytes, platelets, hemoglobin;
– *blood biochemical tests*: CRP, procalcitonin, creatinine, urea, total protein, sodium, potassium, LDG, ferritin, conjugated bilirubin, AST, ALT, troponin I, APTT, glucose, amylase, D-dimer, fibrinogen.

All the data were downloaded from the local health information system (HIS) and manually checked for possible outliers. Off-scale values were set to the bound of an equipment dynamic range. For the restoration of missed values between two sequential tests, the “last observation carried forward” procedure (LOCF) was applied [15]. To compare two cohorts, baseline values for the listed parameters were determined for all the patients.

### Statistical analysis

The analysis was performed based on all the available data from the described cohorts. Sex was coded with values 0 and 1 for females and males respectively. Continuous data were characterized by their medians and MADs due to their non-normality. Categorical data were summarized as proportions. We compared baseline characteristics between the training and validating cohorts using the Mann-Whitney U test for continuous data and the χ^2^ test for categorical data. Cumulative incidence functions were compared with the Gray test. A correlation was estimated with a Spearman’s rank coefficient. All *p*-values were two-sided, all the confidence intervals – 95 percent.

To investigate the association between dynamics of the features and outcomes, a ten days period before an outcome (death or discharge) was analyzed. For every time-varying feature, we fitted a robust linear regression model including a day before an outcome, an outcome itself and their interaction as independent variables, whereas the feature value as a dependent one [16]. Thus, an interaction coefficient sign was used as an indicator whether a feature increase or decrease is more typical in case of expected lethal outcome. Depending on it, either an upper or a lower cut-off value was calculated for every feature. For static features (Age, Sex and BMI), a day term and interaction were excluded from the model, and a cut-off type was selected basing on outcome coefficient sign. When discussing the cut-off level, we didn’t find any unambiguous arguments for prioritizing either sensitivity or specificity, that is why this level were chosen to minimize their average.

Basing on the conventional Bayesian approach [17], the resulting partial sensitivity and specificity of a feature were combined into a feature informativeness index (FII) as follows (see Supplement A for details):

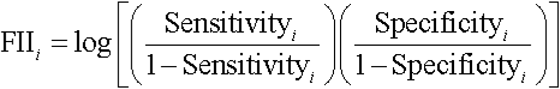

Also for every particular feature we estimated a median prediction time range (defined as a time between the first true positive prediction for a patient and their death). Afterwards, a subset of features with the best informativeness and/or prediction range was selected to be combined into the proposed risk score (see section Results below). The score may be represented as a weighted sum of individual predictions, made with a particular feature. The rounded values of FII’s were used as weights in the score. The detailed description of rationales underlying the described procedure may be found in Supplement A.

For the validation of the proposed score we compared its sensitivity, specificity and prediction range between training and validating cohorts. Also, a testing rate influence on the score was examined by means of formation of a separate patients group (36 cases), in which every score component was intentionally tested not only according to a doctor daily decision but also for ensuring the testing period to be no more than 3 days. A quantile-quantile plot (QQ-plot) was used for the comparison of score values distributions in this group and in the control group in which testing rate was chosen in a conventional fashion (according to the doctors daily decisions only). The control group was chosen with a propensity score matching algorithm basing on baseline score components values.

For the analysis of the relationship between a patient score and their individual mortality risk we fit logistic regression with terms including wave, maximal score during hospitalization and their interaction. The second and the third terms were included for the evaluation of the relationship robustness to the score application conditions. The obtained with this model expected death:discharge odds were split into 5 sub-ranges (grades) with boundary values 1:100, 1:25, 1:5 and 1:1 reflecting very low, low, average, high and very high risks correspondingly.

Finally, we used the small amount of available data related to various cytokines tested occasionally since the beginning of the pandemic for the analysis of correlations between the score and the cytokines levels. The correlations were estimated not only for the coinciding moments but also for a few time lags between the score calculation and the cytokines testing. It is worth noting that these cytokines were measures with a research purpose only and were not used for a treatment strategy improvement.

## Results

### Comparison of cohort

The baseline cohort characteristics of patients analyzed in the study are given in Table 1. As we can see, a noticeable part of patients from the considered cohorts have never experienced such tests as Ferritin, Procalcitonin, Troponin I and LDG. Moreover, a separate analysis showed that these tests were taken mostly for senior patients and patients in a severe status. Hence, to avoid the omitted variable bias, they were excluded from the further analysis. Comparing other features, we may conclude that even in cases with detected statistically significant difference, the clinical difference was considerably low. As a result, the differences between cumulative incidence functions for both events (death and discharge) were statistically insignificant (see Figure 1). However, we should note a slightly worse condition of the 2^nd^ wave patients on average.

**Table 1.** Characteristics of the training (Wave 1) and validation (Wave 2) cohorts

| Test | Patients with known baseline values |  |  | Patients tested at least once |  |  | Average testing period (days) |  |  | Baseline values (median±MAD)* |  |  |
| --- | --- | --- | --- | --- | --- | --- | --- | --- | --- | --- | --- | --- |
|  | I | II | *p* | 1 | 2 | *p* | I | II | *p* | I | II | *p* |
| Age | NA | NA | NA | NA | NA | NA | NA | NA | NA | 58±13.3 | 60±14.8 | <0.001 |
| Sex | NA | NA | NA | NA | NA | NA | NA | NA | NA | F: 752 (55,7%)<br>M: 597 (44,3%) | F: 752 (51,8%)<br>M: 701 (48,2%) | 0,029 |
| BMI | NA | NA | NA | NA | NA | NA | NA | NA | NA | 28.6±5.34 | 28.3±5.12 | 0,227 |
| ALT | 1338<br>(99.2%) | 1446<br>(99.5%) | 0,385 | 1345<br>(99.7%) | 1452<br>(99.9%) | 0,20 | 5,3 | 6,1 | <0.001 | 30±17 | 30.2±17.2 | 0,496 |
| Amylase | 1318<br>(97.7%) | 1440<br>(99.1%) | 0,005 | 1328<br>(98.4%) | 1447<br>(99.6%) | 0,003 | 7,5 | 7,8 | <0.001 | 57±22.2 | 57±23.7 | 0,547 |
| APTT | 1316<br>(97.6%) | 1416<br>(97.5%) | 0,961 | 1326<br>(98.3%) | 1424<br>(98%) | 0,579 | 6,9 | 7,6 | <0.001 | 31.1±4.74 | 33±5.19 | <0.001 |
| AST | 1338<br>(99.2%) | 1446<br>(99.5%) | 0,385 | 1345<br>(99.7%) | 1452<br>(99.9%) | 0,202 | 5,3 | 6,1 | <0.001 | 36±15.6 | 37±16.3 | 0,220 |
| Conj. bilirubin | 1325<br>(98.2%) | 1441<br>(99.2%) | 0,038 | 1334<br>(98.9%) | 1447<br>(99.6%) | 0,046 | 6,9 | 7,3 | <0.001 | 2.5±1.19 | 2.5±1.19 | 0,969 |
| Creatinine | 1339<br>(99.3%) | 1447<br>(99.6%) | 0,367 | 1346<br>(99.8%) | 1453<br>(100%) | 0,111 | 5,4 | 5 | 0,014 | 0.088±0.019 | 0.089±0.019 | 0,159 |
| CRP | 1339<br>(99.3%) | 1445<br>(99.4%) | 0,693 | 1348<br>(99.9%) | 1453<br>(100%) | 0,481 | 2,4 | 2,2 | <0.001 | 46±54 | 54.1±54.6 | 0,001 |
| D-dimer | 1041<br>(77.2%) | 1263<br>(86.9%) | <0.001 | 1241<br>(92%) | 1403<br>(96.6%) | <0.001 | 4,9 | 4,3 | <0.001 | 577±391 | 519±371 | 0,008 |
| Ferritin | 799<br>(59.2%) | 1003<br>(69%) | <0.001 | 976<br>(72.3%) | 1269<br>(87.3%) | <0.001 | 7,2 | 6 | <0.001 | 304±259 | 454±372 | <0.001 |
| Fibrinogen | 1304<br>(96.7%) | 1410<br>(97%) | 0,644 | 1319<br>(97.8%) | 1422<br>(97.9%) | 0,897 | 6,6 | 8,8 | <0.001 | 5.1±1.59 | 5.2±1.5 | <0.001 |
| Glucose | 1332<br>(98.7%) | 1445<br>(99.4%) | 0,073 | 1340<br>(99.3%) | 1451<br>(99.9%) | 0,033 | 5,9 | 5,9 | 0,356 | 6.47±0.98 | 6.8±1.3 | <0.001 |
| Hemoglobin | 1348<br>(99.9%) | 1447<br>(99.6%) | 0,157 | 1349<br>(100%) | 1453<br>(100%) | 1,000 | 2,6 | 2,5 | 0,001 | 139±14.8 | 137±15.6 | 0,010 |
| LDG | 740<br>(54.9%) | 797<br>(54.9%) | 1,000 | 962<br>(71.3%) | 1035<br>(71.2%) | 0,967 | 6,4 | 6,3 | 0,146 | 268±93.4 | 265±93.4 | 0,965 |
| Lymphocytes | 1348<br>(99.9%) | 1447<br>(99.6%) | 0,157 | 1349<br>(100%) | 1453<br>(100%) | 1,000 | 2,6 | 2,5 | 0,001 | 1.2±0.519 | 1.1±0.45 | 0,000 |
| Monocytes | 1348<br>(99.9%) | 1447<br>(99.6%) | 0,157 | 1349<br>(100%) | 1453<br>(100%) | 1,000 | 2,6 | 2,5 | 0,001 | 0.45±0.193 | 0.44±0.2 | 0,315 |
| Neutrophils | 1346<br>(99.8%) | 1446<br>(99.5%) | 0,405 | 1348<br>(99.9%) | 1453<br>(100%) | 0,481 | 2,6 | 2,5 | 0,001 | 3.77±1.79 | 4.21±2.2 | <0.001 |
| Platelets | 1348<br>(99.9%) | 1447<br>(99.6%) | 0,157 | 1349<br>(100%) | 1453<br>(100%) | 1,000 | 2,6 | 2,5 | 0,001 | 203±65.2 | 214±69.2 | <0.001 |
| Potassium | 1340<br>(99.3%) | 1445<br>(99.4%) | 0,878 | 1346<br>(99.8%) | 1452<br>(99.9%) | 0,357 | 5,4 | 5 | 0,008 | 4±0.445 | 4±0.45 | 0,019 |
| Procalcitonin | 892<br>(66.1%) | 529<br>(36.4%) | <0.001 | 1031<br>(76.4%) | 665<br>(45.8%) | 0,000 | 5,2 | 7,2 | <0.001 | 0.11±0.058 | 0.12±0.06 | 0,015 |
| Sodium | 1340<br>(99.3%) | 1445<br>(99.4%) | 0,878 | 1346<br>(99.8%) | 1452<br>(99.9%) | 0,357 | 5,5 | 5,3 | 0,149 | 139±3.26 | 138±3.11 | <0.001 |
| Toponin I | 765<br>(56.7%) | 356<br>(24.5%) | 0,000 | 910<br>(67.5%) | 486<br>(33.4%) | 0,000 | 8,2 | 8,2 | 0,671 | 2±2.97 | 3±02.97 | 0,010 |
| Total protein | 1333<br>(98.8%) | 1443<br>(99.3%) | 0,240 | 1342<br>(99.5%) | 1449<br>(99.7%) | 0,372 | 7,3 | 7,5 | 0,012 | 72±5.93 | 71±5.93 | <0.001 |
| Urea | 1331<br>(98.7%) | 1443<br>(99.3%) | 0,127 | 1339<br>(99.3%) | 1449<br>(99.7%) | 0,107 | 6,4 | 5,8 | 0,004 | 5.1±1.93 | 5.1±1.93 | 0,668 |
| WBC | 1348<br>(99.9%) | 1447<br>(99.6%) | 0,157 | 1349<br>(100%) | 1453<br>(100%) | 1,000 | 2,6 | 2,5 | 0,001 | 5.7±2.09 | 6.03±2.28 | 0,001 |

\* Except sex, for which the data is represented with percentages.

### Estimation of clinical laboratory parameters dynamics before outcome

The analysis of features behavior before an outcome allowed highlighting two groups of features with significant association between lethal outcome and increased/decreased value:

- Increased: Age, Amylase, APTT, AST, Conj. bilirubin, Creatinine, CRP, D-dimer, Glucose, Neutrophils, Sex, Sodium, Urea, WBC;
- Decreased: BMI, Hemoglobin, Lymphocytes, Monocytes, Platelets, Total protein.

For the remaining features their associations with an outcomes were insignificant, which caused there exclusion from the further analysis. The full summaries of regression results, as well as plots of features variation before an outcome are given in Supplement B.

### Evaluation of features informativeness and prediction range

Basing on the first wave data, for every patient we found the worst detected value of every feature (highest or lowest, depending on its behavior given in Table 2). These values allowed fitting a set of partial prediction models (one per feature) and derive cut-offs, providing the minimum of FPR and FNR arithmetic mean – (FPR+FNR)/2. Thus for every feature we calculated its partial specificity, sensitivity, precision, informativeness index FII (according to the procedure, described in Supplement A) and prediction range. The summary of this procedure is given in Table 2 and depicted in Figure 2.

**Table 2.** Partial prediction effectiveness of the features

| Feature | Sensitivity | Specificity | Precision | Threshold | Threshold type | FII | Median prediction range, days |
| --- | --- | --- | --- | --- | --- | --- | --- |
| Age | 0,59 | 0,83 | 0,18 | 68 | Upper | 1,95 | 9,5 |
| Amylase | 0,68 | 0,89 | 0,28 | 104 | Upper | 2,82 | 5 |
| APTT | 0,93 | 0,88 | 0,32 | 42,4 | Upper | 4,49 | 7 |
| AST | 0,80 | 0,83 | 0,23 | 83 | Upper | 2,98 | 5 |
| BMI | 0,41 | 0,71 | 0,08 | 25,95 | Lower | 0,50 | 9 |
| Conj. bilirubin | 0,88 | 0,71 | 0,16 | 3,3 | Upper | 2,87 | 9 |
| Creatinine | 0,85 | 0,91 | 0,39 | 0,123 | Upper | 4,11 | 6 |
| CRP | 0,84 | 0,81 | 0,22 | 146 | Upper | 3,11 | 8,5 |
| D-dimer | 0,86 | 0,86 | 0,28 | 2149 | Upper | 3,64 | 8 |
| Glucose | 0,91 | 0,82 | 0,25 | 8,9 | Upper | 3,89 | 8 |
| Hemoglobin | 0,84 | 0,82 | 0,23 | 114,5 | Lower | 3,14 | 7 |
| Lymphocytes | 0,85 | 0,84 | 0,26 | 0,7 | Lower | 3,44 | 9 |
| Monocytes | 0,54 | 0,78 | 0,14 | 0,23 | Lower | 1,47 | 7,5 |
| Neutrophils | 0,84 | 0,91 | 0,36 | 12,23 | Upper | 3,91 | 6,5 |
| Platelets | 0,58 | 0,87 | 0,22 | 129 | Lower | 2,20 | 5 |
| Sex | 0,53 | 0,56 | 0,07 | 0,5 | Upper | 0,38 | 10 |
| Sodium | 0,74 | 0,92 | 0,38 | 145,1 | Upper | 3,52 | 6 |
| Total protein | 0,98 | 0,89 | 0,35 | 61 | Lower | 5,73 | 7 |
| Urea | 0,95 | 0,92 | 0,44 | 10,9 | Upper | 5,43 | 8 |
| WBC | 0,86 | 0,88 | 0,32 | 13,5 | Upper | 3,87 | 7 |

**Figure 2.**
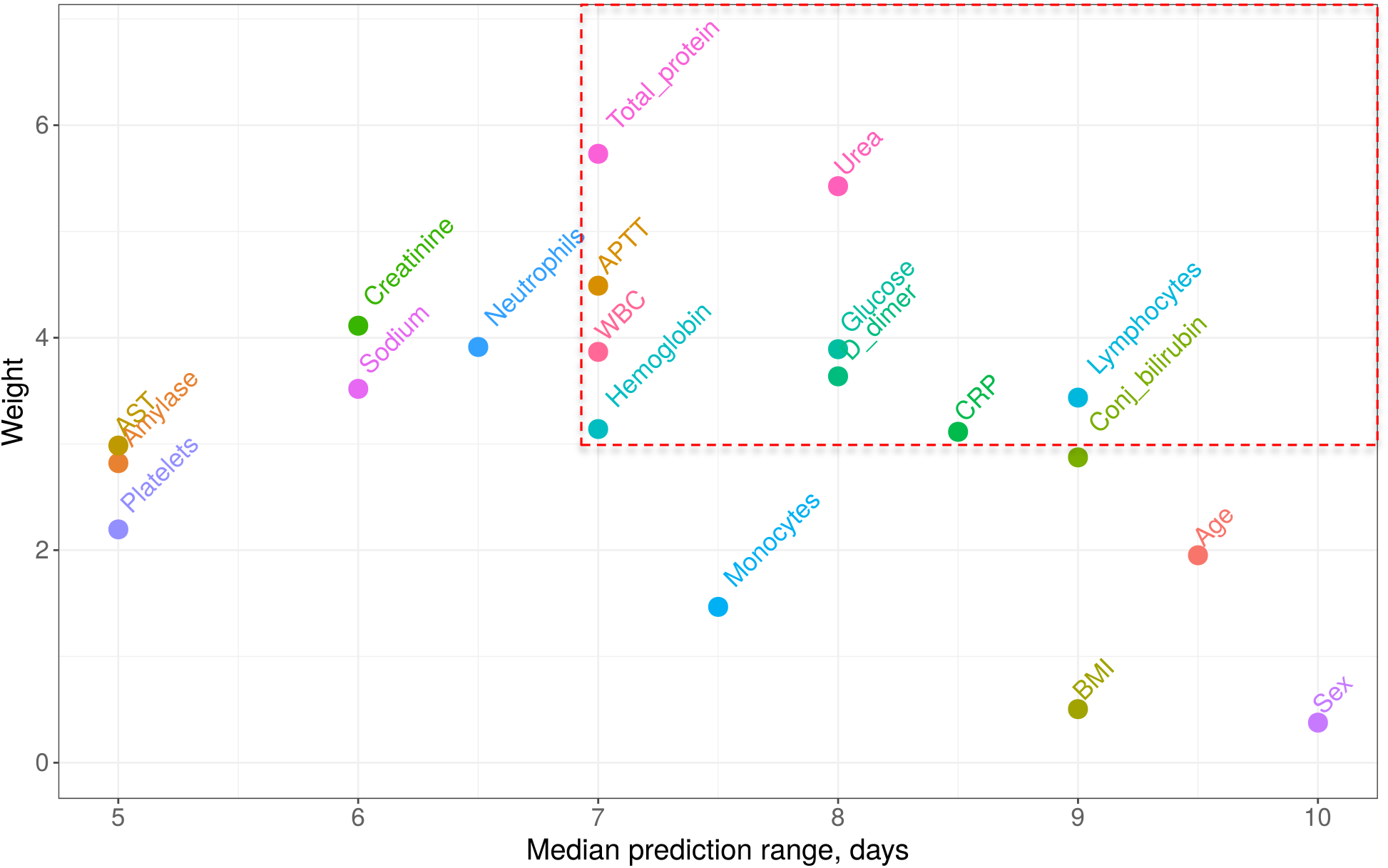
Features informativeness indexes vs. median prediction range. Features with prediction range ≥7 days range and weight ≥3 were accepted as the components of the proposed score (circled with red dashed line)

### Prognostic score statement and its validation

From all the features listed in Table 3 we heuristically extracted only those, which median prediction range was at least 7 days and which information index was at least 3 (circled with red dashed line in figure 2). This resulting set of features with corresponding weights calculated as rounded FIIs are given in Table 3.

**Table 3.** The proposed score components

| Feature | Threshold | Weight |
| --- | --- | --- |
| APTT | > 42 sec | 4 |
| CRP | > 146 mg/L | 3 |
| D-dimer | > 2149 mg/L | 4 |
| Glucose | > 9 mmol/L | 4 |
| Hemoglobin | < 115 g/L | 3 |
| Lymphocytes | < $0,7 \cdot 10^9/L$ | 3 |
| Total protein | < 61 g/L | 6 |
| Urea | > 11 mmol/L | 5 |
| WBC | > $13,5 \cdot 10^9/L$ | 4 |
| Total score: |  | 36 max |

Average score values per day among all the included in this research patients for both waves see in Figure 3. In Figure 4 we represent an obtained sensitivity/specificity trade-off for the proposed prognostic index as well as its mean prediction range dependence on a chosen threshold level.

**Figure 3.**
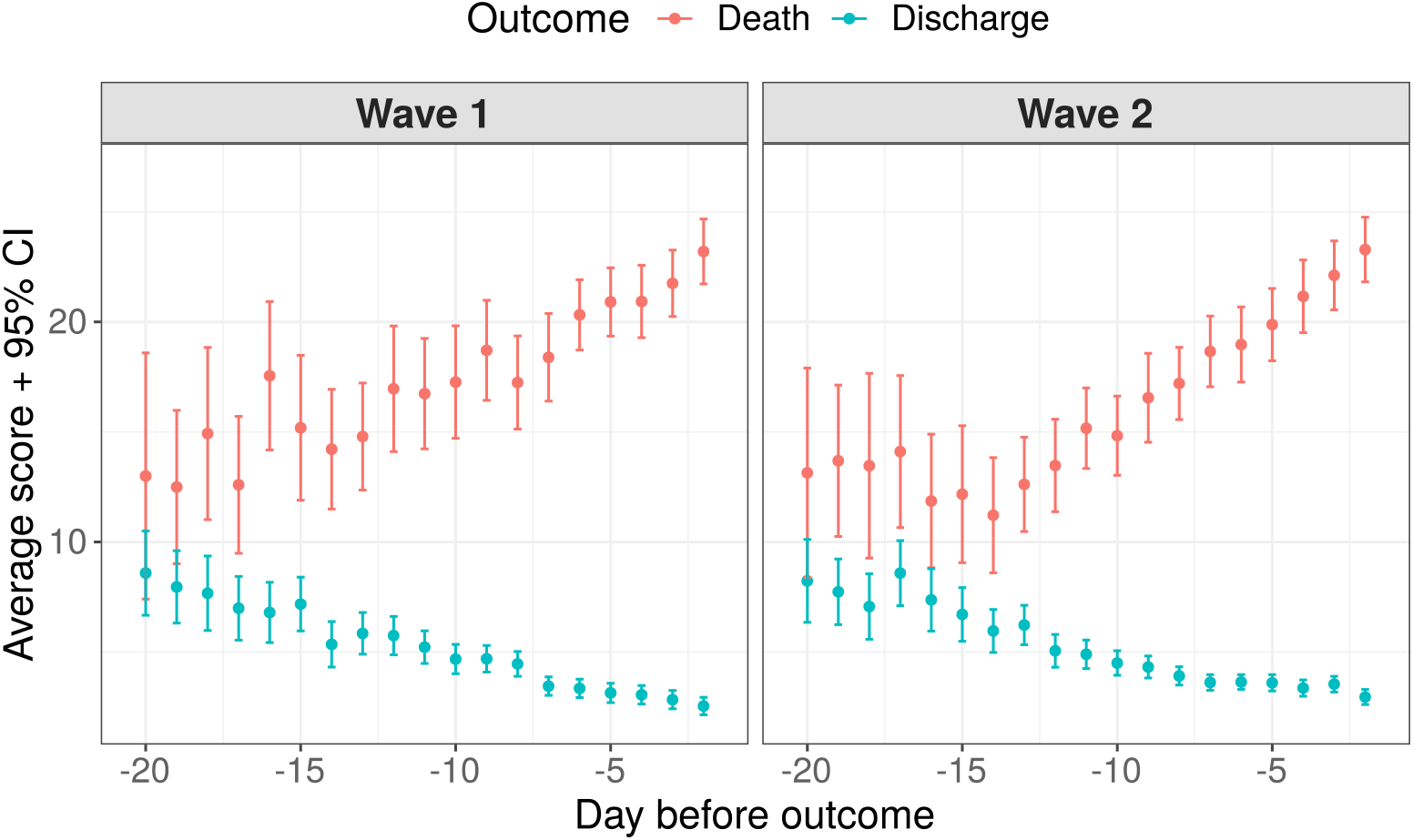
Average score variation within 3 weeks before outcome: points represent mean score values at particular days, error bars – 95% confidence intervals for the mean values

**Figure 4.**
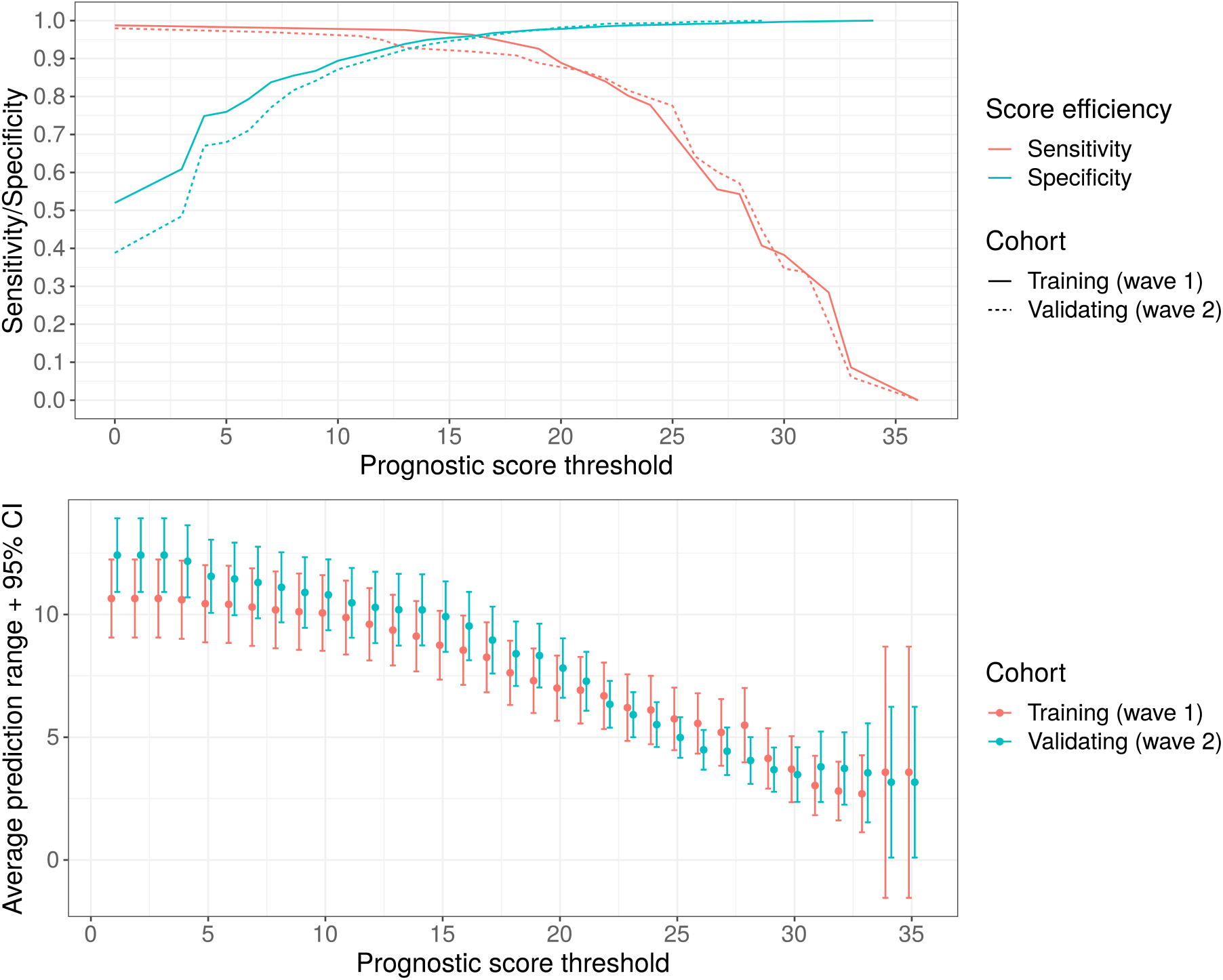
Results of score validation: a) sensitivity/specificity trade-off for various threshold levels; b) Prediction range dependency on a chosen threshold level

Figure 5 represents a QQ-plot with the score quartiles comparison between samples with an experimental (regular) and a conventional testing rate. A fitted linear regression line is depicted for the estimation of potential systematic biases introduced with a testing rate practice.

**Figure 5.**
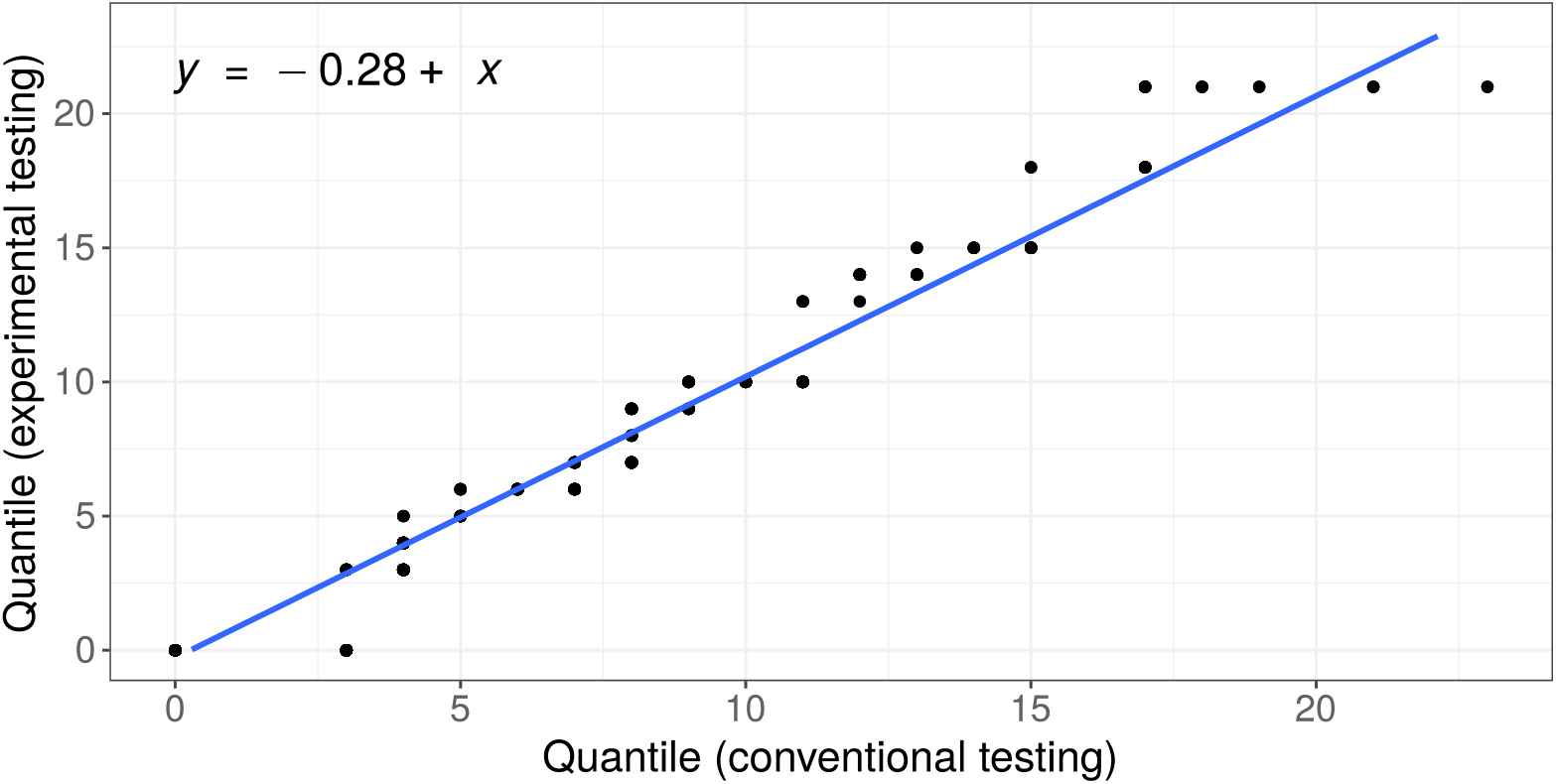
The comparison of score values distributions obtained by means of conventional (according to daily doctor decisions) and experimental (at least once per three days) testing strategies

### Choice of score grades according to a patient individual risk

For a simplification of patient individual risk estimation we introduced the following four risk grades: low, medium, high and very high. Basing on a training cohort data we adjusted the score bounds for their matching with expected death/discharge odds 1:4, 1:1 and 3:1. These bounds as well as odds confidence intervals for both cohorts are given in Table 5 and Figure 6.

**Table 5.** Risk grades according to the prognostic score

| Score range | Expected death/discharge odds | Risk grade |
| --- | --- | --- |
| <9 | < 1:100 | Very low |
| [9, 14) | 1:100 – 1:25 | Low |
| [14, 19) | 1:25– 1:5 | Average |
| [19, 24) | 1:5 – 1:1 | High |
| $\geq 24$ | > 1:1 | Very high |

**Figure 6.**
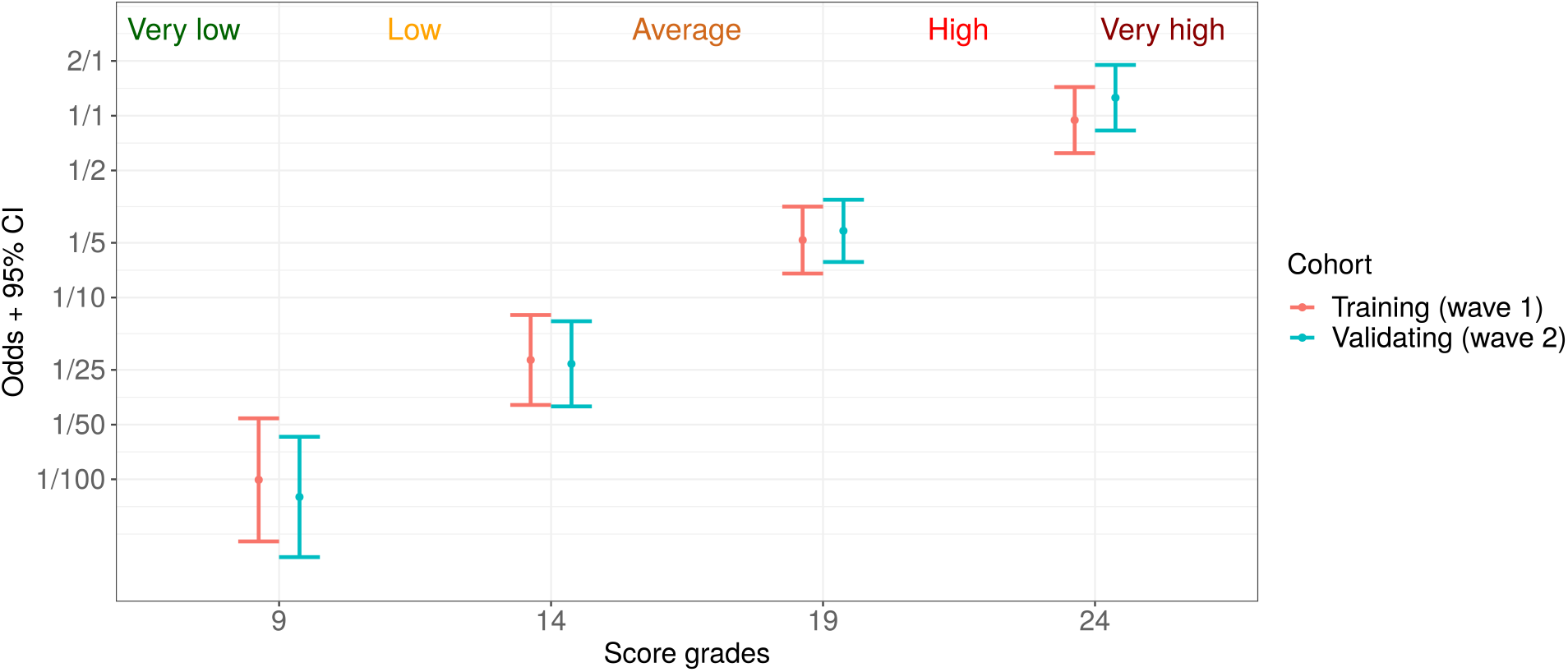
Five risk grades based on proposed prognostic score: very low (death:discharge odds < 1:100), low (1:100 – 1:25), average (1:25– 1:5), high (1:5 – 1:1) and very high (> 1:1)

### Prognostic score and cytokines landscape

In Figure 7 we represent the result of analysis of associations between various cytokines levels and the proposed score. As one can see, the score value has significant correlation with IL-1RA (positive) and IL-1α (negative) measured 5-7 days before. For IL-6 and IL-8 the largest positive correlation was detected for the 3-5 days prior period.

**Figure 7.**
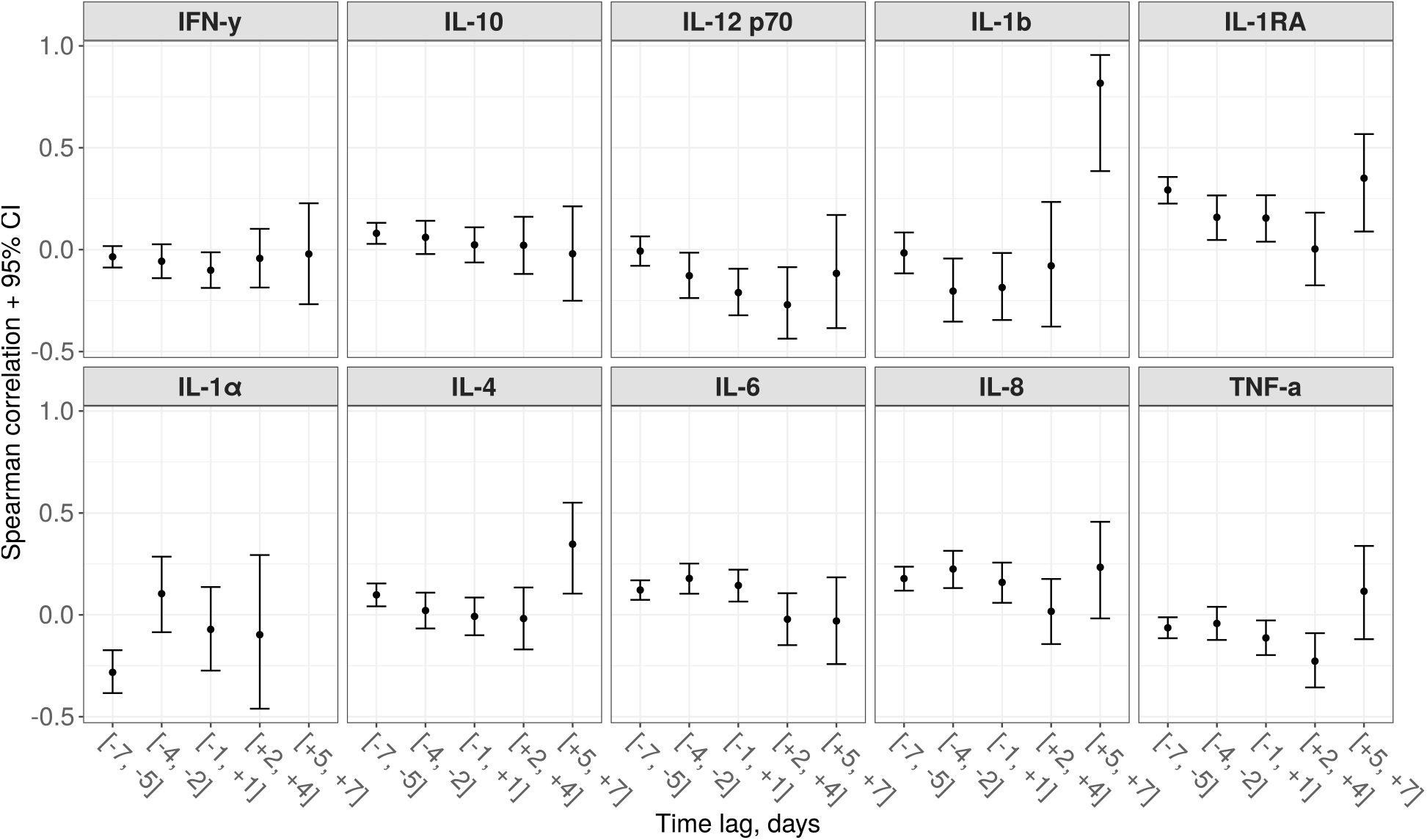
Correlations between the proposed score and 10 various cytokines for different time lags (a negative lag represents cytokins testing preceding to the score calculation, while a positive one – on a contrary, succeeding)

### Score application examples

In this subsection three cases are given for illustrating a routine application of the proposed prognostic score. For all the patients COVID-19 pneumonia was diagnosed with a PCR test and a CT scan on hospital admission. The score variations for the considered cases are given in Figure 8.

**Figure 8.**
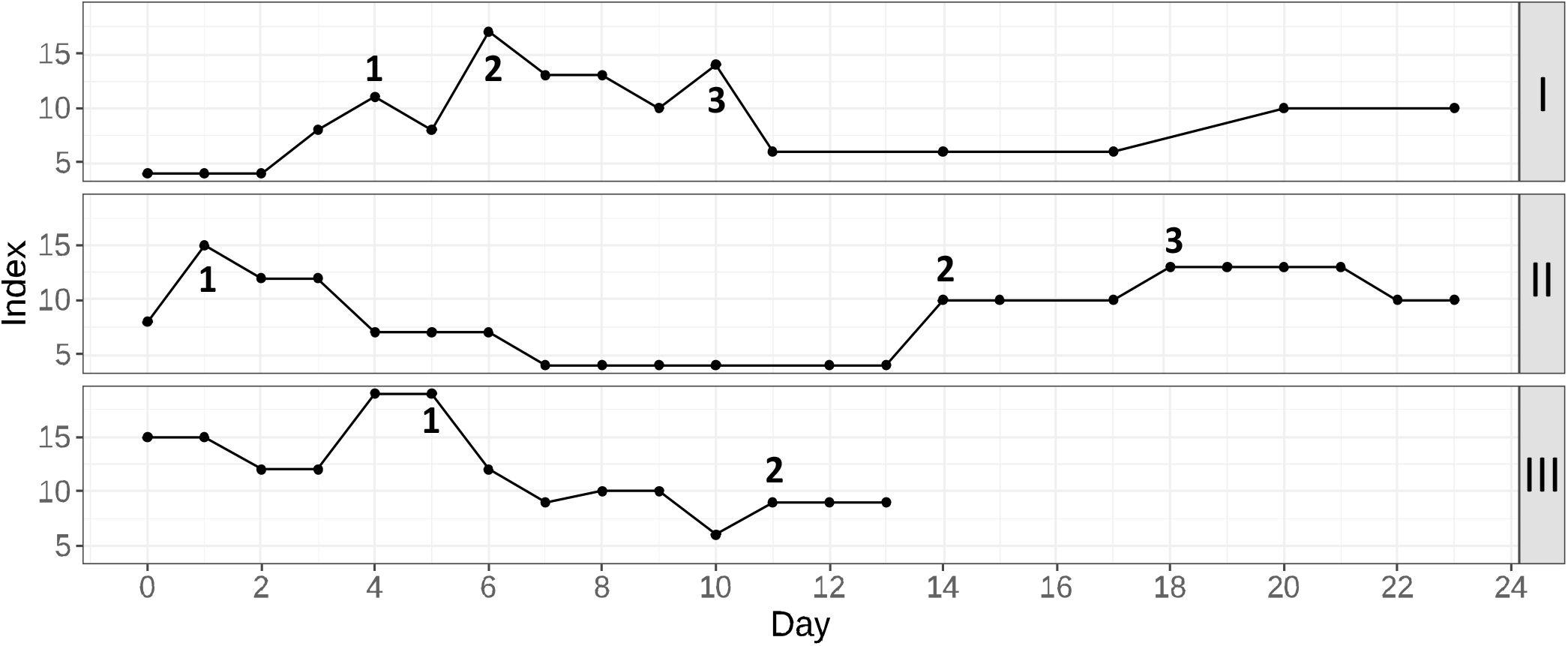
Three examples of the proposed prognostic score application in a routine clinical practice. *Patient I*: 1 – start of anticytokine+antibacterial therapy, 2 – plasma exchange session, 3 – non-invasive ventilation in the general observation unit; *Patient II*: 1 – the moment of the beginning of glucocorticosteroid therapy, 2 – discovery of the hematoma, 3 -a discontinuation of anticoagulant therapy; *Patient III*: 1 – start of anticytokine+antibacterial therapy, 2 – start of additional antimycotic therapy.

Patient I (upper part of the figure) was admitted to the hospital with a moderate respiratory failure (room-air SpO2 – 94%). Within the first four days of hospitalization, the patient’s condition gradually worsened which expressed in increasing of lymphopenia, WBC, neutrophils, CRP and ferritin levels. Moreover a glucose levels was poorly controlled. Therefore therapy with tocilizumab was undertaken in combination with antibiotic therapy (point 1). This resulted in a temporal general improvement which included all the key laboratory parameters except for lymphocytes and ferritin. Room-air SpO2 remained at 89-90%, and the second CT scan was performed. A significant negative trend was found, and it was decided to transfer the patient to the intensive care unit (ICU) for a plasma exchange session and a high-flow oxygen therapy (point 2). After this treatment, there was a significant positive dynamics of clinical and laboratory parameters. The patient was transferred to the general observation unit, where the mitigation of respiratory failure was continued using non-invasive ventilation (point 3). Soon the patient’s condition stabilized and he could be discharged home.

Patient II (middle part of the figure) was hospitalized with a concomitant uncontrolled arterial hypertension and type 2 diabetes. She had severe respiratory failure (room-air SpO2 – 88-90%), extreme fatigue and unsteady gait. For correction of respiratory failure, glucocorticosteroid therapy was undertaken (point 1), followed by the infusion of the immune anti-SARS-CoV-2 plasma and antibiotic therapy (due to the increased procalcitonin level). Then, clinical and laboratory improvement occured. The target blood pressure was reached and the oxygen saturation recovered to 94%. However, during a preparation to a discharge on the 13th day of hospitalization, the sporadic growth of the score was detected. The discharge process was delayed and soon an unsteady gait as well as deterioration of respiratory function, and increasing weakness. An examination of the patient showed decrease in hemoglobin, accompanied with CRP elevation. During CT with contrast enhancement (point 2), the formation of a rounded area of a fluid accumulation with smooth, clear contours in the left pectoralis minor was found. The lesion spread totally from the left axillary region to the level IV-V of the sternocostal joint, with signs of unsharp infiltration of the surrounding tissue. The formation of a hematoma with an element of secondary inflammation was diagnosed. A discontinuation of anticoagulant therapy, and maintenance therapy (point 3) were performed. When the patient’s condition stabilized and improved, she was discharged.

Patient III (lower part of the figure) was admitted with a respiratory failure that required a glucocorticosteroid therapy from the first day of hospitalization. After a short-term condition improvement, a critical deterioration was detected by means of the proposed prognostic score. Due to the fact that there was no progression of a respiratory failure, and the patient did not make any complaints, it was difficult to assess his condition by means of daily physical examination. Given the high risk of poor outcome, anticytokine therapy with a JAK-kinase inhibitor in combination with antibiotic therapy was prescribed (point 1). Subsequently, a dramatic improvement in the patient’s condition was observed. On the 11th day of hospitalization, the additional antimycotic therapy was required due to the colonization of respiratory tract and oral cavity with Candida spp. (point 2). Upon completion of the therapy course, the patient was discharged.

### Analysis of the most noticeable prediction failures

For the analysis of the score application limitations we have manually checked the most noticeable cases with a prediction failure. In this dataset 3 deaths occurred with a score value less than 9 points (“very low” risk grade). In all these cases the deaths were caused by an extremely rapid deterioration: in two cases, associated with the acute cardiovascular failure (for patients aged 60-65 years and 80-85 years), in the third case – with the development of subdural hematoma of the right hemisphere of the brain (for patient aged 90-95 years). Thus, there were not enough time for performing the laboratory tests and updating the score.

Also, we consider 12 cases in which patients had score values more than 30, but survived and were discharged. In 5 of 12 patients, the observed decompensation was caused by prehospital therapy (in 2 cases antibacterial therapy resulted in pseudomembranous colitis, in 3 cases intensive anticoagulant therapy – bleeding and formation of hematomas). In 4 of 12 patients, a severe course of COVID-19 was observed with significant impairment of respiratory function and the need to prescribe anticytokine therapy and non-invasive ventilation. This therapy successfully delayed the development of acute respiratory distress syndrome. Subsequently, the patients required the appointment of antibiotic therapy due to secondary bacterial complications after anticytokine therapy, which proved to be effective. In 3 of 12 patients, the high score values reflected the progression of chronic diseases (stenosing cancer of sigmoid colon, breast cancer and polycystic kidney disease). At the same time, a course COVID-19 was mild, so, after effective management of chronic diseases, the patients were successfully discharged.

## Discussions

Almost all the selected features were mentioned earlier as prognostic factors for COVID-19 patients. Thus, in research [18] urea together with creatinine reflects the state of kidney function at the time of COVID-19 illness, indicating a higher risk of severe infection in patients with chronic kidney disease (CKD) or acute kidney injury (AKI). In addition to indicating the current renal pathology and nephrotoxicity of the applied therapy, urea is one of the signs of catabolism in critically ill patients. The relationship between urea and catabolism has previously been demonstrated in patients with extensive trauma due to persistent muscle catabolism/rhabdomyolysis [19]. Later the predictive potential of urea was demonstrated not only in patients with polytrauma, but also in severe patients with other conditions [20]. Increase in aspartate aminotrasferase (AST) levels in patients with COVID-19 indicates a possible contribution of an active inflammatory process to urea metabolism due to the involvement of muscle tissue. Inflammatory destruction of muscle tissue can also act as a marker of the severity of the SARS-CoV-2 infection.

The concomitant appearance of abnormalities of conjugated bilirubin, AST and ALT may also indicate an underestimated role of liver dysfunction in pathogenesis of COVID-19 within hepatorenal syndrome [21]. Analogously, the dynamic decrease in total protein by the time of lethal outcome can also be considered an indirect reflection of both an increasing renal dysfunction during COVID-19 in patients with CKD, and a sign of “minor” hepatorenal syndrome [22], [23].

High levels of C-reactive protein (CRP) and ferritin are frequently considered as a criteria for the so-called cytokine storm [24]. Being the main feature of SARS-CoV-2 infection, this phenomena also includes a concomitant increase in procalcitonin [25], which indicates a high concentration of pro-inflammatory cytokines with a simultaneous low level of IFN-gamma [26]. In this situation we have to deal with a difficult differential diagnosis of secondary bacterial/fungal superinfection in COVID-19 patients. The difficulty is also associated with neutrophilic leukocytosis, which also develops during widely practiced corticosteroid therapy.

A low level of lymphocytes in the early stages of infection is considered as a criterion for subsequent severity of the infection course [27]. Cytokine storm together with a lung injury are associated with a decrease in circulating lymphocytes both as a result of their direct depletion and their infiltration of the affected lung tissue. Hence, the undulating course of COVID-19 may be associated with new episodes of a lymphocyte level decrease [28].

A disturbance of glucose metabolism in patients with COVID-19 and its relationship with the severity of the course of infection may be the reflection of: i) poorly controlled diabetes in patients at the moment of hospitalization; ii) a side effect of glucocorticosteriod therapy during infection; iii) more rarely – de novo development of diabetes as a consequence of SARS-CoV-2 infection [29]. Hence, persistently elevated glucose levels in COVID-19 patients may be associated with a higher risk of secondary bacterial superinfection.

A decrease in hemoglobin during a COVID-19 has alse been discussed as a predictor of an unfavorable course of infection previously [30]. In the early stages of the disease, it can be associated with a high concentration of circulating IL-6 and the risk of a cytokine storm. In the later stages of the disease, hemoglobin may be an indirect sign of a violation of coagulation with a bleeding episode.

Finally, coagulation abnormalities in COVID-19 patients are almost unavoidable satellites of this infection leading to difficult-to-manage complications, such as venous thrombosis and thromboembolism. Also bleeding episodes due to a massive anticoagulant therapy may occur in these patients [31]. Particularly, a gastrointestinal bleeding due to a steroid and an anticytokine therapy in patients with pre-existing pathologies of mucosa may be distinguished. For this reason, regular monitoring of both D-dimer and APTT in patients is the most effective strategy for controlling the balance between anticoagulant and anti-inflammatory therapies.

Thus, the mentioned features accepted as the score components cover almost all the peculiarities of COVID-19 pathogenesis. The comparison of a training and a validating cohorts shows, that despite seasonal effects, change of paradigm in COVID-19 treatment and difference between baseline feature values, the proposed approach shows a high accuracy and robustness. Thus, for both cohorts it was shown that 11-19 points cut-offs provides score sensitivity and specificity over 90% with an expected prediction range >7 days (see Figure 4). Moreover, the differences in death:discharge odds between the considered cohorts for the selected risk grades were shown to be statistically insignificant (see Figure 6).

### Study limitations

The study has the following limitations. Firstly, it was a single-center study with a temporal validation. Secondly, a few features that showed the efficiency in previous studies (such as ferritin, LDH, procalcitonin and troponin), were excluded from the analysis due to insufficiency of data. Also, laboratory tests for various features were done asynchronously and with different updating periods. This indicates different degrees of the score components relevance at the time of its calculation. For this reason, we assume that the score distribution will depend on the rate of analyses sampling in each particular hospital. Finally, the score doesn’t contain any components reflecting a patient respiratory function. Thus, an objective estimation of a patient condition may be performed only in combination with oxygenation parameters.

## Conclusion

An extensive testing of the score during the second wave of COVID-19 has demonstrated its attractiveness for application by a medical staff due to a precision in representing the current patient state and interpretability. Besides, it was shown, that the proposed score may be used as a tool for automatic detection of patients with a dangerous deterioration and thus help to reduce the burden on a staff during high-workload periods.

## Data Availability

The datasets analysed in this study are available from First Pavlov State Medical University officials on reasonable request.

## Acknowledgements

The authors would like to acknowledge medical staff of Pavlov University who participated in the testing of the developed score, in particular Irina V. Shlyk, Alexey A. Afanasiev, Alexandra V. Novikova and Alexandra A. Lebedeva. We would also like to show our gratitude to Maria D. Vladovskaya and Alexander M. Ginzburg for their valuable advices on working in the local health information system.

## Declarations

### Ethics approval

The study was performed in compliance with the World Medical Association Declaration of Helsinki on Ethical Principles for Medical Research Involving Human Subjects, and was reviewed by a local Review Board.

