## Supplement A for "A precise score for the regular monitoring of COVID-19 patients condition validated within the first two waves of the pandemic"

### Supplement A. A heuristic procedure for score components weights assignment

To optimize the score components weights, we assume that a set of $N$ various laboratory tests is available. Each test $i$ produces result $X_i$ which may be either $\hat{D}_i$ in case of predicted death or $\hat{S}_i$ when successful discharge is predicted. Let $\alpha_i$ and $\beta_i$ be test $i$ marginal false positive and false negative error rates correspondingly. To draw a final decision basing on all the tests results, the following *a posteriori* (AP) probability ratio may be written:

$$AP\left(X_1 \dots X_N\right) = \frac{\Pr\left\{D \mid X_1 \dots X_N\right\}}{\Pr\left\{S \mid X_1 \dots X_N\right\}} = \frac{\Pr\left\{D\right\}\Pr\left\{X_1 \dots X_N \mid D\right\}}{\Pr\left\{S\right\}\Pr\left\{X_1 \dots X_N \mid S\right\}} \qquad \text{(A.1)}$$

If a preliminary correlation analysis was performed and the given set of tests has a reasonably small mutual dependence, the following approximation of expression (A.1) may be proposed:

$$AP\left(X_1 X_2 \dots X_N\right) \approx \frac{\Pr\left\{D\right\}\prod_{i=1}^{N}\Pr\left\{X_i \mid D\right\}}{\Pr\left\{S\right\}\prod_{i=1}^{S}\Pr\left\{X_i \mid D\right\}} \qquad \text{(A.2)}$$

Let's split the products in the nominator and the denominator in (A.2) into two sub-products depending on marginal decisions results:

$$\frac{\Pr\left\{D\right\}\prod_{i=1}^{N}\Pr\left\{X_i \mid D\right\}}{\Pr\left\{S\right\}\prod_{i=1}^{S}\Pr\left\{X_i \mid D\right\}} = \frac{\Pr\left\{D\right\}\prod_{X_i=\hat{D}_i}\Pr\left\{X_i \mid D\right\}\prod_{X_i=\hat{S}i}\Pr\left\{X_i \mid D\right\}}{\Pr\left\{S\right\}\prod_{X_i=\hat{D}_i}\Pr\left\{X_i \mid S\right\}\prod_{X_i=\hat{S}_i}\Pr\left\{X_i \mid S\right\}} =$$

$$= \frac{\Pr\left\{D\right\}\prod_{X_i=\hat{D}_i}\Pr\left\{\hat{D}_i \mid D\right\}\prod_{X_i=\hat{S}_i}\Pr\left\{\hat{S}_i \mid D\right\}}{\Pr\left\{S\right\}\prod_{X_i=\hat{D}_i}\Pr\left\{\hat{D}_i \mid S\right\}\prod_{X_i=\hat{S}_i}\Pr\left\{\hat{S}_i \mid S\right\}} \qquad \text{(A.3)}$$

The substitution of $\alpha_i$ and $\beta_i$ instead of conditional probabilities in (A.3) results in the following expression:

$$\frac{\Pr\{D\}\prod_{X_i=\hat{D}_i}\Pr\{\hat{D}_i\mid D\}\prod_{X_i=\hat{S}_i}\Pr\{\hat{S}_i\mid D\}}{\Pr\{S\}\prod_{X_i=\hat{D}_i}\Pr\{\hat{D}_i\mid S\}\prod_{X_i=\hat{S}_i}\Pr\{\hat{S}_i\mid S\}}=\frac{\Pr\{D\}\prod_{X_i=\hat{D}_i}(1-\beta_i)\prod_{X_i=\hat{S}_i}\beta_i}{\Pr\{S\}\prod_{X_i=\hat{D}_i}\alpha_i\prod_{X_i=\hat{S}_i}(1-\alpha_i)}=$$

$$=\frac{\Pr\{D\}}{\Pr\{S\}}\prod_{X_i=\hat{D}_i}\left(\frac{1-\beta_i}{\alpha_i}\right)\prod_{X_i=\hat{S}_i}\left(\frac{\beta_i}{1-\alpha_i}\right)$$

The latter expression may be rewritten using indicator functions:

$$\frac{\Pr\{D\}}{\Pr\{S\}}\prod_{X_i=\hat{D}_i}\left(\frac{1-\beta_i}{\alpha_i}\right)\prod_{X_i=\hat{S}_i}\left(\frac{\beta_i}{1-\alpha_i}\right)=\frac{\Pr\{D\}}{\Pr\{S\}}\prod_i\left(\frac{1-\beta_i}{\alpha_i}\right)^{I\{X_i=\hat{D}_i\}}\left(\frac{\beta_i}{1-\alpha_i}\right)^{I\{X_i=\hat{S}_i\}}=$$

$$=\frac{\Pr\{D\}}{\Pr\{S\}}\prod_i\left(\frac{1-\beta_i}{\alpha_i}\right)^{I\{X_i=\hat{D}_i\}}\left(\frac{\beta_i}{1-\alpha_i}\right)^{1-I\{X_i=\hat{D}_i\}}=\frac{\Pr\{D\}}{\Pr\{S\}}\prod_i\left(\frac{\beta_i}{1-\alpha_i}\right)\left[\left(\frac{1-\beta_i}{\alpha_i}\right)\left(\frac{1-\alpha_i}{\beta_i}\right)\right]^{I\{X_i=\hat{D}_i\}}$$

Applying a logarithmic function to the AP ratio gives:

$$\log AP(X_1\ldots X_N)=\log\frac{\Pr\{D\}}{\Pr\{S\}}+\sum_i\log\left(\frac{\beta_i}{1-\alpha_i}\right)+\sum_i I\{X_i=\hat{D}_i\}\log\left[\left(\frac{1-\beta_i}{\alpha_i}\right)\left(\frac{1-\alpha_i}{\beta_i}\right)\right] \tag{A.4}$$

A conventional classification approach consists in comparing $\log AP(X_1\ldots X_N)$ with a threshold chosen according to a required sensitivity/specificity trade-off. As we can see, the first two components of the sum in (A.4) don't depend on values $X_1\ldots X_N$. Thus, they may be omitted without loss of prediction efficiency. The resulting score function will be written then as follows:

$$Score=\sum_i I\{X_i=\hat{D}_i\}\omega_i\,.$$

Here:

$$\omega_i = \left[ \left( \frac{1-\beta_i}{\alpha_i} \right) \left( \frac{1-\alpha_i}{\beta_i} \right) \right]$$

In the above equation [.] denotes a rounding procedure.
