## Supplement B for "A precise score for the regular monitoring of COVID-19 patients condition validated within the first two waves of the pandemic"

### Supplement B. Analysis of patients features dynamics before outcome

Table B. Robust regression coefficients for considered features

| Feature | Intercept | Day before outcome | Lethal outcome | Interaction | *p*-value | Threshold_type |
|---|---|---|---|---|---|---|
| Age | 55,561 | - | 17,24004 | - | 1E-06 | Upper |
| ALT | 62,013 | 1,792829 | -6,73908 | -0,45107 | 0,277375 | Lower |
| Amylase | 65,096 | 0,401711 | 7,440098 | 1,897953 | 2,8E-06 | Upper |
| APTT | 30,931 | -0,15563 | 23,48463 | 1,847344 | 7,92E-63 | Upper |
| AST | 43,385 | -0,04688 | 28,82057 | 2,487885 | 1,68E-14 | Upper |
| BMI | 29,069 | - | -0,85677 | - | 0,206482 | Lower |
| Conj_bilirubin | 2,272 | -0,04336 | 1,795281 | 0,123558 | 7,79E-12 | Upper |
| Creatinine | 0,081 | -0,00053 | 0,074193 | 0,00656 | 3E-83 | Upper |
| CRP | 16,446 | -3,84408 | 113,1996 | 5,847141 | 3,39E-29 | Upper |
| D_dimer | 875,320 | -9,03413 | 3386,114 | 210,0928 | 1,67E-51 | Upper |
| Ferritin | 420,134 | -1,10278 | 422,527 | 1,478446 | 0,886502 | Upper |
| Fibrinogen | 4,840 | -0,04959 | 0,083639 | -0,01128 | 0,656259 | Lower |
| Glucose | 6,361 | -0,05777 | 3,12614 | 0,055615 | 0,043945 | Upper |
| Hemoglobin | 131,372 | -0,23233 | -28,2304 | -0,59104 | 0,003274 | Lower |
| LDG | 246,656 | -8,00863 | 428,0746 | 27,90315 | 5,88E-41 | Upper |
| Lymphocytes | 1,737 | 0,050565 | -0,98658 | -0,05384 | 1,45E-18 | Lower |
| Monocytes | 0,608 | 0,011606 | -0,11799 | -0,01207 | 1,64E-05 | Lower |
| Neutrophils | 3,822 | -0,10006 | 8,764783 | 0,46006 | 2,86E-47 | Upper |
| Platelets | 313,745 | 7,587005 | -143,614 | -13,7101 | 2,41E-35 | Lower |
| Potassium | 4,477 | 0,036149 | 0,232668 | 0,007987 | 0,259182 | Upper |
| Procalcitonin | 0,098 | -0,00445 | 1,39404 | 0,112736 | 0 | Upper |
| Sex | 0,451 | - | 0,086531 | - | 0,521291 | Upper |
| Sodium | 141,021 | 0,185574 | 4,88449 | 0,244052 | 6,48E-07 | Upper |
| Toponin I | 0,002 | -0,00021 | 0,056907 | 0,004587 | 7,1E-179 | Upper |
| Total_protein | 66,428 | -0,21607 | -13,8551 | -0,34804 | 0,001211 | Lower |
| Urea | 5,291 | -0,07739 | 16,97786 | 1,107044 | 2,9E-112 | Upper |
| WBC | 6,462 | -0,0296 | 7,680757 | 0,387139 | 5,59E-28 | Upper |

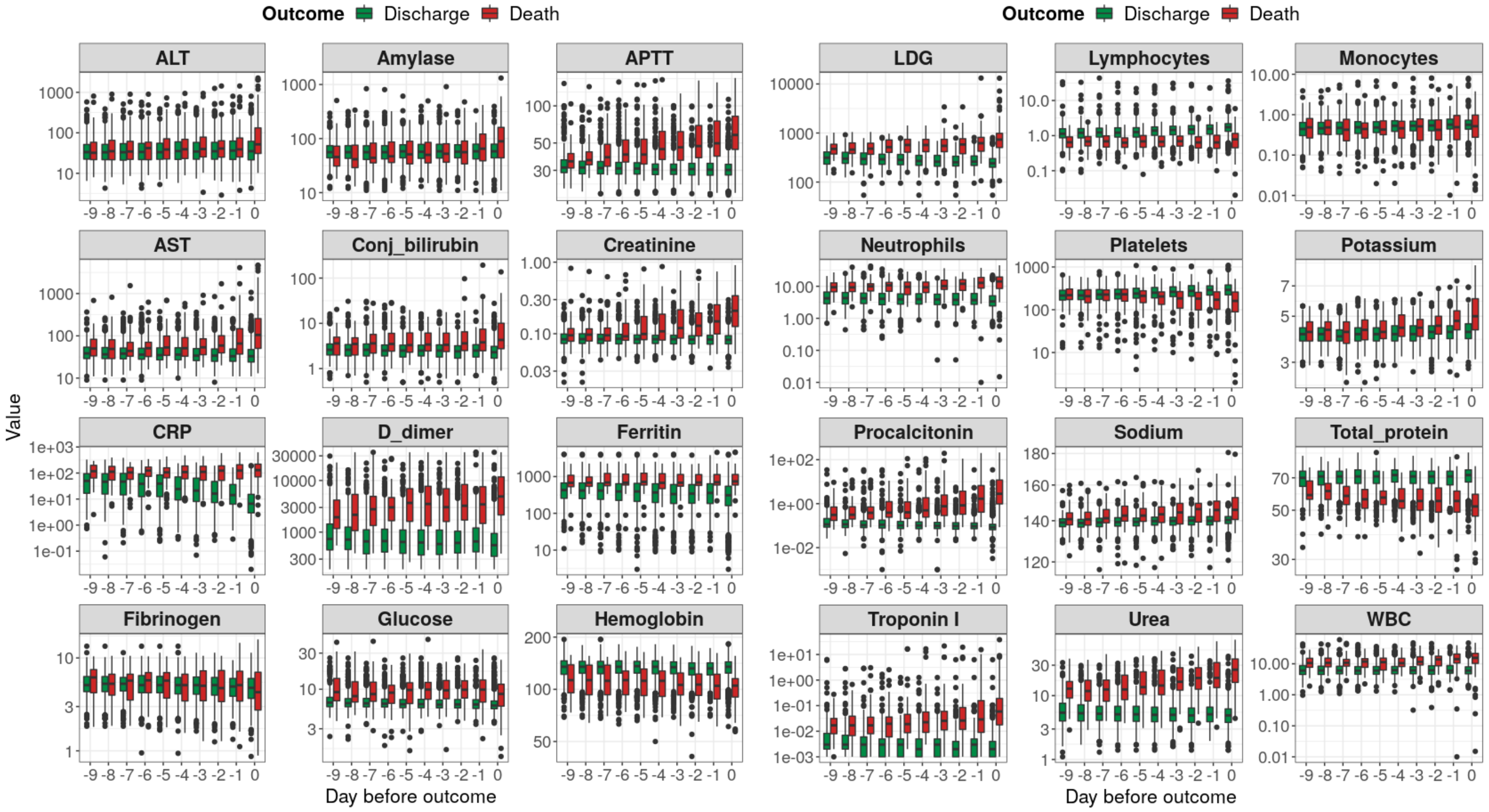


Figure B. Time-varying features behavior before outcome (training cohort – wave 1)
